# Targeted WASHmobile Program for Climate Anticipatory Action to Reduce Anemia during Extreme Weather Events in the Democratic Republic of the Congo

**DOI:** 10.64898/2026.11.19.26368050

**Authors:** Hannah Greene, Kelly Endres, Presence Sanvura, Jean-Claude Bisimwa, Camille Williams, Jamie Perin, Justin Bengehya, Jean Claude Kulondwa, Alves Namunesha, Willy Felicien, Ghislain Maheshe, Justin Cirhuza Cikomola, Alain Mwishingo, Lucien Bisimwa, Anna Dimitrova, Laura Caulfield, Christine Marie George

## Abstract

**Background:** Evidence-informed anticipatory action is critical to mitigate the health impacts of extreme weather events. This randomized controlled trial (RCT) investigated the impact of the WASHmobile hygiene and water treatment program on anemia during extreme weather in eastern Democratic Republic of the Congo (DRC).

**Methods:** The WASHmobile RCT enrolled participants in Bukavu, DRC, from November 2021 to December 2022 (NCT05166850). Anemia was classified by WHO thresholds. CHIRPS data was used to calculate Standardized Precipitation Indices (SPI).

**Findings:** Among 1,208 participants, 68% (822/1208) were anemic. Extreme wet (SPI-1≥2.0) (odds ratio (OR) 2.29; 95% CI 1.03, 5.11), severe wet (1.5≤SPI-1<2) (OR 3.25; CI 1.53, 6.93), and moderate drought conditions (−1.5 < SPI-1 ≤ −1.0) (OR = 2.09; CI 1.28, 3.40) over the prior month were associated with significantly higher odds of anemia. In the standard arm, anemia was significantly higher during extreme wet conditions compared to near-normal conditions (78% vs. 62%, p=0.0496), whereas a non-significant decrease was observed in the WASHmobile arm (64% vs. 69%, p=0.55). Consistent with this, significant effect modification was observed between extreme wet conditions and WASHmobile delivery on anemia prevalence (multiplicative interaction p=0.044). These findings informed the design of the WASHmobile anticipatory action framework where pre-defined climate triggers activate an early warning system for anemia prevention.

**Interpretation:** Extreme and severe wet conditions and moderate drought were associated with higher odds of anemia. WASHmobile presents a potential climate-resilient intervention to reduce anemia during extreme weather.

**Funding:** Wellcome Trust, UK government Foreign, Commonwealth & Development Office, and the National Institutes of Health.

## INTRODUCTION

Extreme weather events such as droughts and floods can heighten anemia risk by increasing outbreaks of diarrheal^1^ and vector-borne diseases.^2^ Anemia impairs child cognitive development, maternal and birth outcomes, and adult productivity.^3^ Droughts and floods can also substantially diminish crop yields and agricultural productivity, affecting nutritional outcomes. ^4,5^ Extreme weather is particularly deleterious for conflict-prone humanitarian settings such as the Democratic Republic of the Congo (DRC) because food supplies can be highly vulnerable to such disruptions.^6^ In South Kivu, where health outcomes are routinely impeded by political instability, conflict, and a high burden of communicable disease, anemia prevalence is estimated to be 66% for children under 5 years and 51% for women of reproductive age.^7^ The DRC also has the second-highest malaria burden in the world,^8^ and diarrhea outbreaks are among the highest in Africa.^9^ However, studies assessing the health impacts of extreme weather events in the DRC^10^ remain limited, and few have examined the associations between climate and anemia. ^11,12^

Climate anticipatory action, in which pre-developed actions are triggered when a climate indicator crosses a predefined threshold, is increasingly used to reduce the health impacts of extreme weather events, but to our knowledge has not been applied to anemia.^13^ Flooding and drought can each contaminate water supplies and push households towards lower-quality water sources, making water treatment and safe water storage essential.^14^ Hygiene interventions can reduce infectious disease transmission by increasing handwashing with soap and improving handwashing infrastructure during climatic events.^15^ Targeted water, sanitation, and hygiene (WASH) interventions could therefore be integrated into climate-resilient early warning and anticipatory action systems alongside infectious disease surveillance to reduce the infectious disease burden contributing to anemia during extreme weather events.^16^

The WASHmobile program integrates healthcare facility diarrhea surveillance with the delivery of WASH mobile health (mHealth) messaging targeted to populations at high risk of diarrheal diseases.^17^ In our randomized controlled trial in the DRC, WASHmobile significantly reduced diarrhea prevalence, healthcare facility visits for diarrhea, serological responses for cholera, and stunting for children under 5 years.^18^ In this study, we aimed to: (i) assess associations between drought and extreme and severe wet conditions with anemia, (ii) examine the impact of WASHmobile on anemia prevalence during extreme weather, and (iii) integrate these findings into the WASHmobile anticipatory action framework for anemia prevention during extreme weather.

## METHODS

### Study design and setting

We conducted a longitudinal analysis within the WASHmobile cluster-randomized controlled trial (RCT) in Bukavu, South Kivu province, DRC.^18^ This climate analysis was not prespecified in the trial protocol. Bukavu is a densely populated city on Lake Kivu, bordering Rwanda, with an elevation of approximately 1600m, extensive informal settlements, and a history of substantial conflict.^19^ The region typically has a bi-modal rainy season with a main dry season from June to September, and heavy rainfall combined with steep terrain frequently causes flooding and landslides.

Households were randomized to the standard arm or the WASHmobile arm. The standard arm received the standard message from the DRC Ministry of Health for diarrhea patients during healthcare facility discharge, a five-minute WASH message and instruction on the use of oral rehydration solution. The WASHmobile arm received this message and the 12-month WASHmobile mobile health (mHealth) program. The program included weekly automated voice and SMS messages from a doctor at a local hospital promoting handwashing with soap, water treatment, and safe drinking water storage (covering stored water), with quarterly in-person visits. The intervention protocol is described in detail elsewhere.^18^

### Study population

The WASHmobile RCT enrolled the households of diarrhea patients recruited through daily diarrhea surveillance in 115 healthcare facilities (both public and private) in Bukavu from November 22, 2021 to December 14, 2022.^18^ Patients with diarrhea were eligible for enrollment if they had been admitted to a healthcare facility with at least three loose stools in a 24-hour period; did not have a functioning tap in their home, which was typical among those living in informal settlements; had a child under five years of age in the household; had at least one functional mobile phone in the household; planned to continue residing in Bukavu for the next year; and provided a stool sample within 24 hours of enrollment. The household members of each index patient were eligible for participation in the RCT if they shared the same cooking pot as the diarrhea patient, lived in the same home for the prior three nights, and planned to continue residing with the patient for the next 12 months.

This analysis included participants 6 months to 65 years of age (the age range covered by WHO anemia thresholds) who resided in households with GPS data (including elevation data) and had at least one valid hemoglobin measurement. Participant-visits reporting iron supplementation within the previous 2 weeks were excluded (n=54).

All study participants or their guardians provided written informed consent. The trial is registered at ClinicalTrials.gov (NCT05166850) and was approved by the Catholic University of Bukavu (7107) and the Johns Hopkins Bloomberg School of Public Health Institutional Review Board (9848). The funders had no role in data collection or analysis.

### Randomization and masking

The study biostatistician (JP) assigned study randomization (1:1) using R (version 3.3.0), and allocation was revealed to the study coordinator (LB) through a digital database by the study manager (PS).^18^ Separate teams conducted intervention and evaluation activities. Masking of participants and intervention staff after baseline was not possible because intervention components were visible. Hemoglobin was measured with a point-of-care analyzer that gives a device-read result, limiting the potential for assessor bias.

### Participant data

Participant and household demographic data were collected at baseline. Monthly clinical surveillance was conducted to assess self-reported health outcomes. Information on iron supplementation in the previous 2 weeks was also collected.

### Hemoglobin and anemia

A venous blood sample was collected at baseline and 1, 6, and 12 months after enrollment. Hemoglobin concentrations were measured in g/dL using the FaStep Hemoglobin Analyzer, a point-of-care diagnostic tool which quantifies the concentration of hemoglobin in blood (manufacturer: HealthcareTuye, China). Anemia status was categorized based on the WHO guidelines which defined anemia as an elevation-adjusted hemoglobin concentration below 105 g/L for children aged 6-23 months; 110 g/L for 24-59 months; 115 g/L for age 5-11 years; 120 g/L for age 12-14 years; 120 g/L for nonpregnant women aged 15-65 years; and 130 g/L for men aged 15-65 years, with thresholds for mild, moderate, and severe classification within each stratum.^20^ Hemoglobin measurements outside of the clinically plausible range of 40 to 200 g/L were removed from analysis as they suggested invalid measurements (n=4).^21^ Hemoglobin concentrations were adjusted for elevation as recommended by the 2024 WHO guidelines to account for how the expected concentration of hemoglobin in an individual’s blood differs by elevation of residence.^20^

### Precipitation assessment

Daily precipitation data were obtained from the Climate Hazards Group InfraRed Precipitation with Station data (CHIRPS) dataset, which provides rainfall quantities from 1981 onward at 0.05°x0.05° resolution (~5.5 kilometers). CHIRPS blends satellite infrared-based precipitation estimates with weather station observations.^22^ Because weather stations are sparse in the Congo Basin, CHIRPS is considered to be among the most robust datasets for this region.^23^ Household GPS coordinates were used to link each participant-visit to its CHIRPS grid cell.

We characterized precipitation using the standardized precipitation index (SPI), which the World Meteorological Organization recommends for monitoring wet and dry anomalies because it is comparable across locations and seasons.^24^ SPI expresses accumulated precipitation over a specified period relative to the long-term precipitation distribution for the same location and time of year, with positive values indicating wetter-than-normal conditions and negative values indicating drier-than-normal conditions. SPI was calculated separately for each CHIRPS grid cell using monthly precipitation totals from 1990-2024 and the SPI function in the SPEI package in R, with accumulation periods of one month (SPI-1) and three months (SPI-3) preceding each visit date. We used these accumulation periods to encompass acute, infection-related mechanisms of anemia and the longer-term effects of abnormally wet or dry conditions on agriculture, livelihoods, nutrition, disease vectors, and daily activities.^5,6^

SPI values were categorized as severe/extreme drought (SPI ≤ −1.5), moderate drought (−1.5 < SPI ≤ −1.0), near normal (−1.0 < SPI < 1.0), moderate wet (1.0 ≤ SPI < 1.5), severe wet (1.5 ≤ SPI < 2.0), or extreme wet (SPI ≥ 2.0).^25^ Severe and extreme drought were combined because few observations were classified as extreme drought. Each category was compared with near-normal conditions.

### Statistical analysis

To investigate the association between categorical SPI (relative to near-normal conditions) and binary anemia status, we fit logistic regression models using generalized estimating equations (GEE) with an exchangeable working correlation to account for household clustering. We conducted two main analyses. The first analysis assessed associations between SPI and anemia, including all standard arm visits and only baseline WASHmobile arm visits, which preceded in-tervention delivery. Post-baseline WASHmobile visits were excluded as the intervention may have altered WASH behaviors associated with infections and, in turn, anemia. In sensitivity analyses, we stratified models by study arm and sex, and modeled hemoglobin concentration as the outcome.

The second analysis assessed whether these associations differed by study arm. It included all post-baseline visits from both arms, adding a multiplicative interaction term to test effect modification between SPI category and study arm.

Models adjusted for participant-reported age, sex, study arm, and household socioeconomic indicators: household density (residents per sleeping room), a concrete floor (vs. dirt or mud floor, an indicator of wealth in this context), and any household income source. Analyses were conducted in R (version 4.5.2) using the geepack package.

## RESULTS

Anemia status was ascertained from 1,208 participants for 1,539 participant-visits, after 104 participant-visits were removed from this analysis for not meeting inclusion criteria (Figure 1). Among this population, 61% (742/1208) of participants were female, and the average baseline age was 13.9 years (range: 0.5-65) (Table 1).

**Table 1.** Baseline characteristics of participants with hemoglobin data.

| **Characteristic** | N | Overall (N=1208) | Standard (N=701) | WASHmobile (N=507) |
|---|---|---|---|---|
| Age at baseline, years | 1208 | 13.9 ± 12.8 (0.0–65.0) | 13.8 ± 12.8 (0.0–63.0) | 14.1 ± 12.8 (0.5–65.0) |
| **Sex** | 1208 |  |  |  |
| Male |  | 466 (38.6%) | 282 (40.2%) | 184 (36.3%) |
| Female |  | 742 (61.4%) | 419 (59.8%) | 323 (63.7%) |
| Existence of household income source | 1189 | 1111 (93.4%) | 632 (91.5%) | 479 (96.2%) |
| People living in household | 1194 | 7.9 ± 2.7 (2.0–16.0) | 7.9 ± 2.6 (3.0–16.0) | 8.0 ± 2.8 (2.0–15.0) |
| Sleeping rooms | 1194 | 2.5 ± 1.0 (1.0–7.0) | 2.4 ± 0.9 (1.0–5.0) | 2.6 ± 1.1 (1.0–7.0) |
| People per sleeping room | 1194 | 3.6 ± 1.5 (0.7–10.0) | 3.7 ± 1.6 (0.8–10.0) | 3.4 ± 1.4 (0.7–8.0) |
| Concrete floor | 1194 | 520 (43.6%) | 263 (37.8%) | 257 (51.6%) |
| Fever in past 2 weeks | 1028 | 151 (14.7%) | 95 (16.0%) | 56 (12.9%) |
| Fever in past 48 hours | 845 | 99 (11.7%) | 56 (13.9%) | 43 (9.7%) |
| Hospital referral in the prior month | 1039 | 64 (6.2%) | 34 (5.7%) | 30 (6.8%) |
| Hemoglobin, g/L, at first visit | 1208 | 109.4 ± 19.1 (38.2–181.2) | 109.1 ± 19.1 (38.2–181.2) | 109.8 ± 19.0 (40.9–181.0) |
| **Anemia severity at first visit** | 1208 |  |  |  |
| No anemia |  | 455 (37.7%) | 265 (37.8%) | 190 (37.5%) |
| Mild anemia |  | 300 (24.8%) | 173 (24.7%) | 127 (25.0%) |
| Moderate anemia |  | 407 (33.7%) | 237 (33.8%) | 170 (33.5%) |
| Severe anemia |  | 46 (3.8%) | 26 (3.7%) | 20 (3.9%) |
| Any anemia at first visit | 1208 | 753 (62.3%) | 436 (62.2%) | 317 (62.5%) |

The baseline characteristics of the 1,208 participants with hemoglobin data are described as Mean ± SD (Min–Max) for continuous variables, or n (%) for categorical variables. Participants missing covariates (such as income source or household density) were excluded from later models. Hemoglobin levels are adjusted for elevation, and measurements outside of the range considered clinically probable by the WHO (40-200 g/L) were excluded.

**Figure 1.**
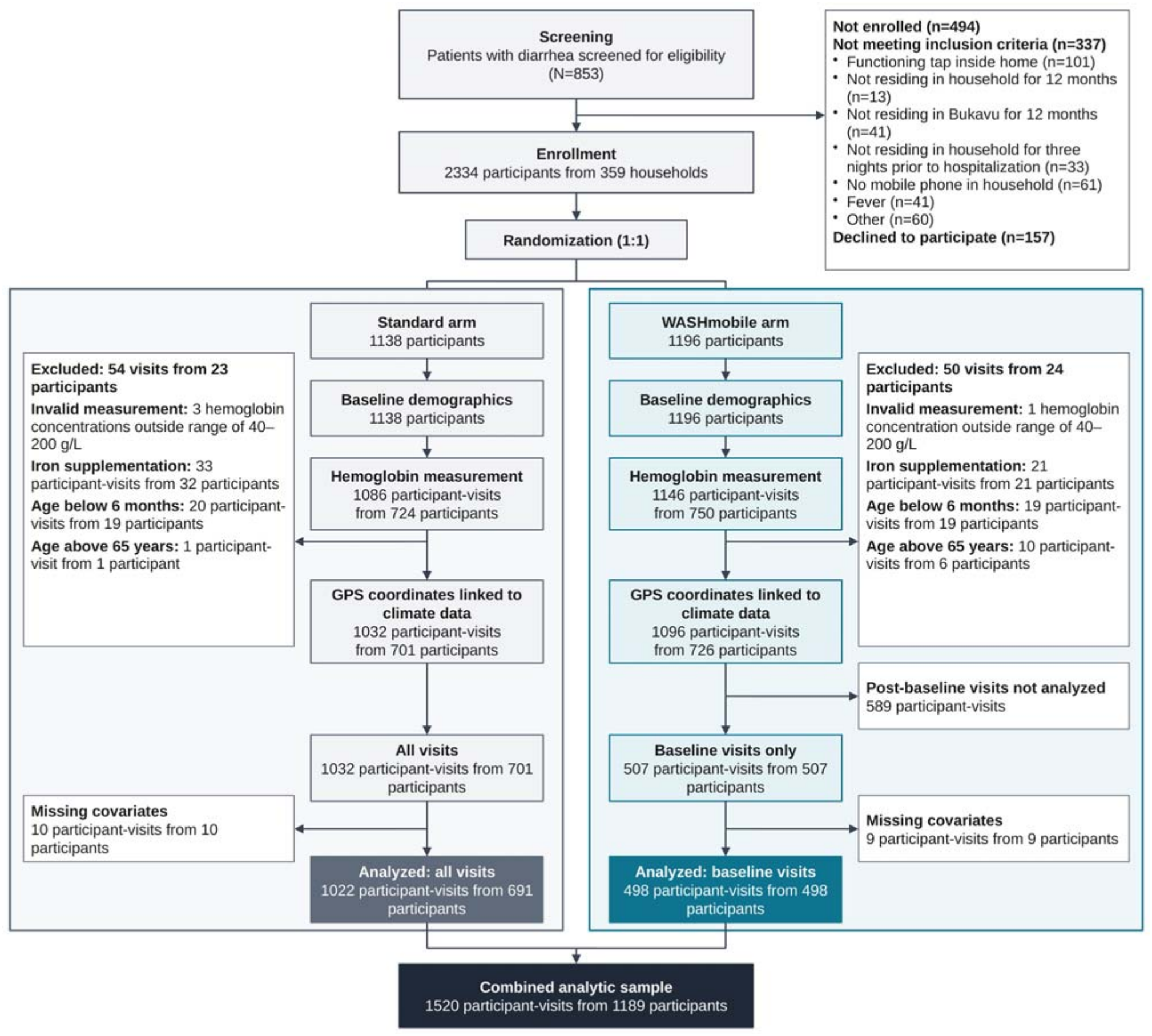
CONSORT diagram illustrating screening, enrollment, and follow-up. Measurements outside of the range considered clinically probable by the WHO (40-200 g/L) were excluded. Participants reporting iron supplementation in the prior two weeks were excluded. Complete-case observations were those with all covariates used in the models (gender, age, household density, floor type, presence of an income source). For the analysis of effect modification by study arm, all participant-visits were included. For the primary analysis of anemia on climate indicators, only baseline visits of the WASHmobile arm were included, because the intervention may have altered behaviors related to infection and subsequent anemia.

Overall, 68% (822/1208) of participants met the WHO definition for elevation-adjusted anemia at one or more visits. The mean hemoglobin concentration was 109.7 g/L. Anemia prevalence was highest among children under 2 years (male: 76.2% (61/80), female: 77.8% (70/90)) and lowest among males aged 15 years or older (56.2% (63/112)) (S1).

Mean monthly rainfall in the study area ranged from 2.1 cm in July to 15.9 cm in November, with a monthly average of 9.97 cm over the study period. Table 2 includes anemia prevalence observed over the study period by SPI category. Of the 1,520 participant-visits included in the first analysis, 916 were classified as near-normal conditions (SPI-1) over the month prior, 73 as severe or extreme drought, 152 as moderate drought, 272 as moderate wet, 35 as severe wet, and 72 as extreme wet conditions.

**Table 2.**
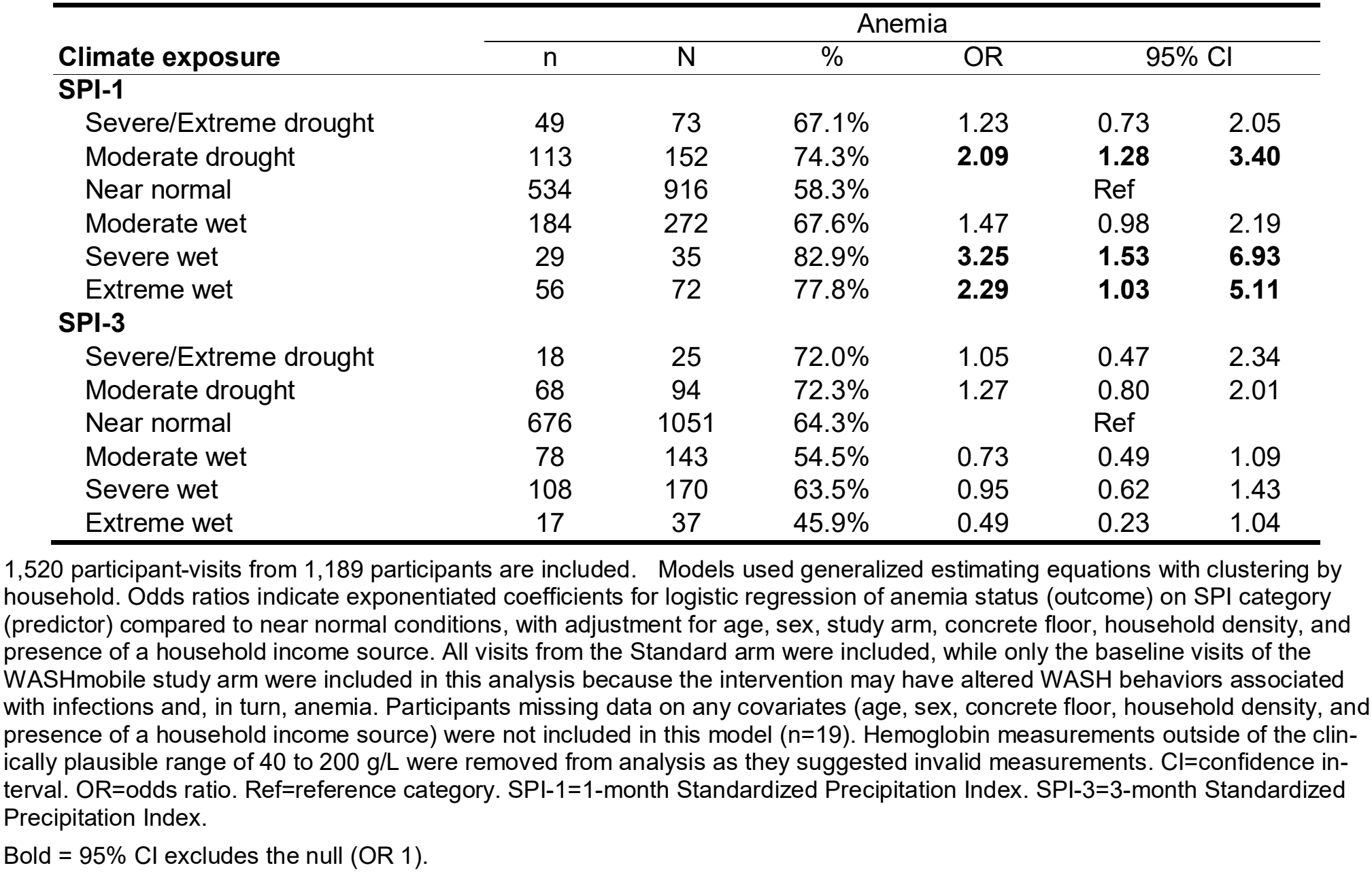
Associations between anemia and SPI.

|  | Anemia |  |  |  |  |  |
|---|---|---|---|---|---|---|
| **Climate exposure** | n | N | % | OR | 95% CI |  |
| **SPI-1** |  |  |  |  |  |  |
| Severe/Extreme drought | 49 | 73 | 67.1% | 1.23 | 0.73 | 2.05 |
| Moderate drought | 113 | 152 | 74.3% | **2.09** | **1.28** | **3.40** |
| Near normal | 534 | 916 | 58.3% | Ref |  |  |
| Moderate wet | 184 | 272 | 67.6% | 1.47 | 0.98 | 2.19 |
| Severe wet | 29 | 35 | 82.9% | **3.25** | **1.53** | **6.93** |
| Extreme wet | 56 | 72 | 77.8% | **2.29** | **1.03** | **5.11** |
| **SPI-3** |  |  |  |  |  |  |
| Severe/Extreme drought | 18 | 25 | 72.0% | 1.05 | 0.47 | 2.34 |
| Moderate drought | 68 | 94 | 72.3% | 1.27 | 0.80 | 2.01 |
| Near normal | 676 | 1051 | 64.3% | Ref |  |  |
| Moderate wet | 78 | 143 | 54.5% | 0.73 | 0.49 | 1.09 |
| Severe wet | 108 | 170 | 63.5% | 0.95 | 0.62 | 1.43 |
| Extreme wet | 17 | 37 | 45.9% | 0.49 | 0.23 | 1.04 |

1,520 participant-visits from 1,189 participants are included. Models used generalized estimating equations with clustering by household. Odds ratios indicate exponentiated coefficients for logistic regression of anemia status (outcome) on SPI category (predictor) compared to near normal conditions, with adjustment for age, sex, study arm, concrete floor, household density, and presence of a household income source. All visits from the Standard arm were included, while only the baseline visits of the WASHmobile study arm were included in this analysis because the intervention may have altered WASH behaviors associated with infections and, in turn, anemia. Participants missing data on any covariates (age, sex, concrete floor, household density, and presence of a household income source) were not included in this model (n=19). Hemoglobin measurements outside of the clin-ically plausible range of 40 to 200 g/L were removed from analysis as they suggested invalid measurements. CI=confidence in-terval. OR=odds ratio. Ref=reference category. SPI-1=1-month Standardized Precipitation Index. SPI-3=3-month Standardized Precipitation Index.
Bold = 95% CI excludes the null (OR 1).

Compared with near-normal conditions, extreme wet (Odds Ratio (OR): 2.29; 95% Confidence Interval (CI): 1.03, 5.11), severe wet (OR: 3.25; CI 1.53, 6.93), and moderate drought conditions (OR: 2.09; CI 1.28, 3.40) in the prior month were associated with higher odds of anemia (Table 2). SPI-3 was not significantly associated with anemia status. For hemoglobin, moderate drought (β −5.7; CI −9.5, −1.8) and severe wet conditions (β −8.9; CI −16.2, −1.6) in the prior month were associated with lower concentrations (S2). Conversely, three months of moderate wet (β 3.17; CI 0.09, 6.24) or severe wet (β 4.37; CI 0.85, 7.90) conditions (SPI-3) were associated with higher concentrations of hemoglobin. Results were similar when the analysis was restricted to the standard arm (S3). The direction of associations between SPI-1 and anemia was consistent across female and male participants (S4).

We found a significant interaction between extreme wet conditions and the study arm on the odds of anemia. In the standard arm, the prevalence of anemia was significantly higher during extreme wet conditions compared to near-normal conditions (OR 3.06, CI 1.00, 9.32; prevalence 78% under extreme wet vs 62% under near-normal conditions), while a non-significant decrease in anemia prevalence was observed within the WASHmobile arm (64% vs. 69%) (Figure 2). Consistent with this, we observed a significant interaction between extreme wet conditions and the study arm on anemia prevalence (multiplicative interaction OR 0.27 (95% CI 0.08, 0.97), p=0.044) (Figure 3 and S5).

**Figure 2.**
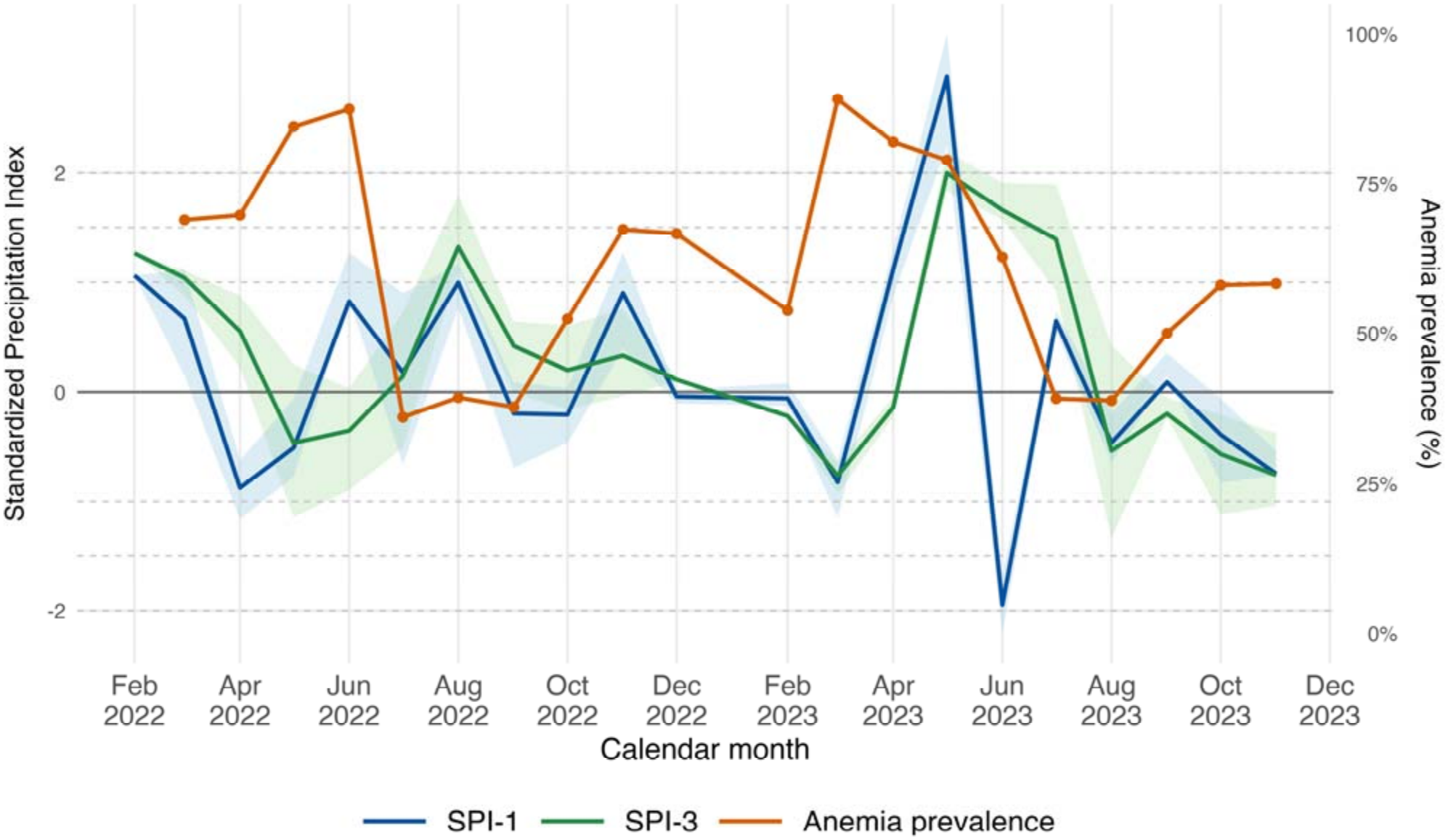
Anemia prevalence and standardized precipitation index over the study period in Bukavu, DRC. Monthly mean one-month or three-month SPI (blue) depicts the deviation of one- or three-month precipitation accumulation relative to the long-term precipitation distribution, with shading indicating the interquartile range of SPI measurements observed across the study area. Anemia prevalence (red) shows the percentage of samples classified as anemic (calculated as number of samples classified as anemic based on WHO thresholds for age and sex after adjusting for altitude, divided by number of samples analyzed).

**Figure 3.**
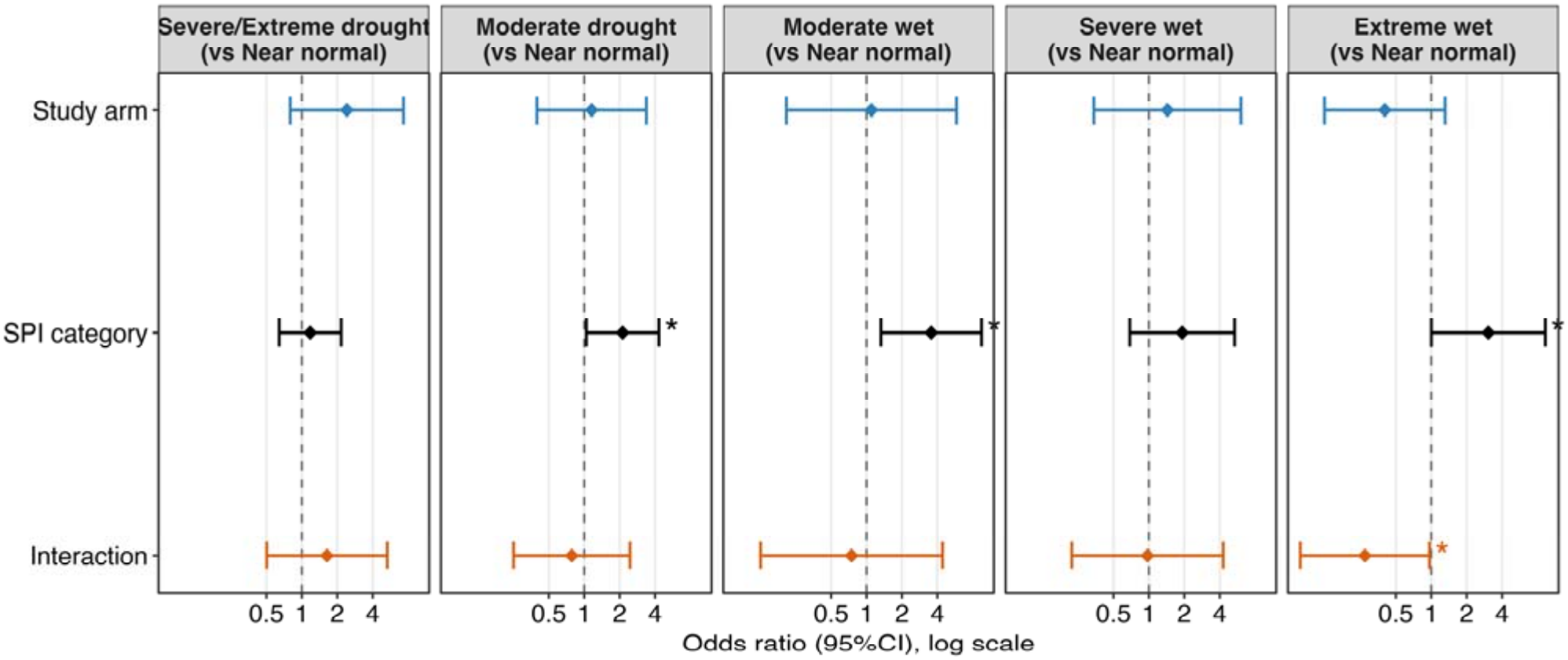
Associations between SPI-1 categories and anemia, with interaction terms for effect modification. For the standard arm, 577 participant-visits from 471 participants are included. For the WASHmobile arm, 577 participant-visits from 474 participants are included. Points represent odds ratios (reported in Appendix Table S5) based on household-clustered logistic GEE models. Coefficients for SPI category (black) compare each SPI category with Near Normal conditions in the reference subgroup (Standard for study arm). The exponentiated coefficients for the effect modifier (WASHmobile arm) within each SPI category are depicted in blue. Exponentiated interaction coefficients (orange) represent the ratio of SPI-category odds ratios for WASHmobile vs Standard. Error bars indicate 95% CI. An asterisk (*) indicates significance at p < 0.05.

### Anticipatory action framework

Based on these findings, we developed the WASHmobile anticipatory action framework for climate-responsive delivery of WASHmobile to reduce anemia during extreme weather in the DRC (Figure 4). In this framework, climate triggers inform deployment of WASHmobile to preemptively reinforce WASH behaviors. The first climate trigger is the one-month SPI, which can be calculated in real time from open-access CHIRPS data. This pre-defined climate trigger aligns with the SPI categories associated with anemia in this study: SPI-1 ≥ 1.5 for wet conditions and SPI-1 ≤ −1.0 for drought. The second climate trigger is based on short-range weather forecasting systems including Google’s Flood Hub and WeatherNext3, which can forecast extreme weather events such as floods days in advance.^26^ Continuous monitoring of climatic conditions can allow for deployment of flood and drought-specific WASHmobile in forecasted areas to reduce infectious disease outbreaks and reduce the anemia burden for the affected population. The WASHmobile anticipatory action framework has three phases: (1) readiness, (2) early action, and (3) response. In the readiness phase, guided by seasonal forecasts and forecasted events such as El Niño, WASHmobile messages are prepared, partnerships are established with mobile network operators for mobile message delivery, community health workers are trained, and supply chains for WASH products are secured. Readiness also includes implementing preventive measures for landslides and crop soil erosion such as terrace farming and barricades to protect residential areas from flooding, promoting covering stored water in the community for malaria prevention, distributing insecticide-treated bed nets, and administering preventative chemoprophylaxis for malaria.^5,14^ In the early action phase, once the threshold for a climatic trigger is crossed, climate-specific WASHmobile messages are deployed (Figure 5). Actions triggered during extreme wet conditions could include emphasis on chlorine treatment of water and handwashing with soap, vigilance for vector habitats (e.g. covering stored water).^14^ Early action in response to periods of drought could include vouchers for chlorine tablets and promotion of alternative water sources for handwashing only (e.g. river water, lake water).^14^ In the third response phase, continued surveillance for outcomes such as anemia, malaria, waterborne disease, and child nutritional status (e.g. MUAC) guides WASHmobile messaging intensity to affected and food-insecure households. This anticipatory action framework was developed in conjunction with a similar framework for cholera and other diarrhea (Endres et al, submitted).

**Figure 4.**
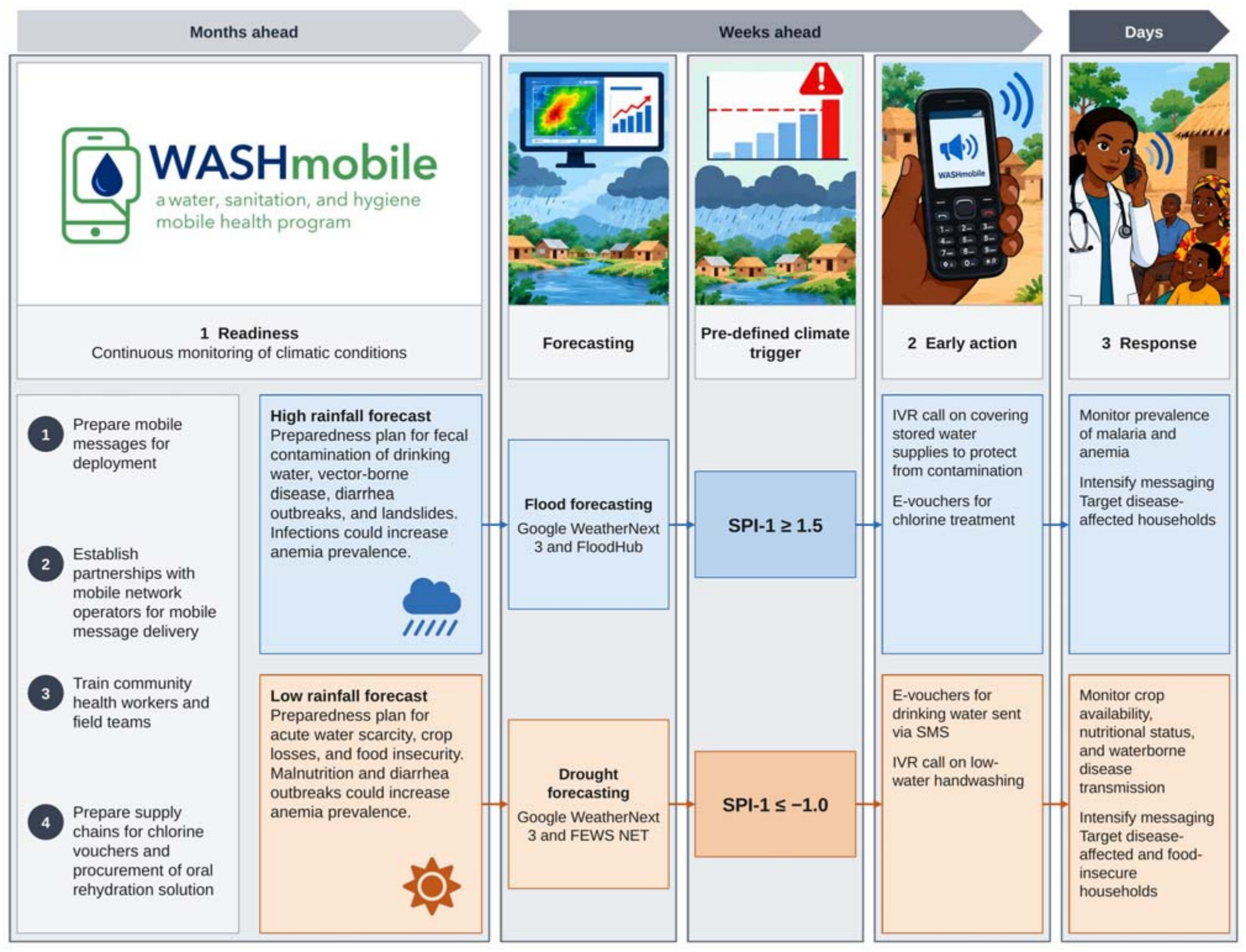
WASHmobile Framework for anticipatory action to reduce anemia during extreme weather in the DRC. The WASHmobile anticipatory action framework has three phases: (1) readiness, (2) early action, and (3) response phases. In this framework, pre-defined climate triggers inform deployment of WASHmobile to preemptively reinforce WASH behaviors. The first climate trigger is the one-month SPI, which can be calculated in real time from open-access CHIRPS data. The second climate trigger is based on short-range weather forecasting systems including Google’s FloodHub and WeatherNext3, which can forecast extreme weather events such as floods days in advance.^26^ Continuous monitoring of climatic conditions can allow for deployment of flood and drought-specific WASHmobile in forecasted areas to reduce infectious disease outbreaks and reduce the anemia burden for the affected population.

**Figure 5.**
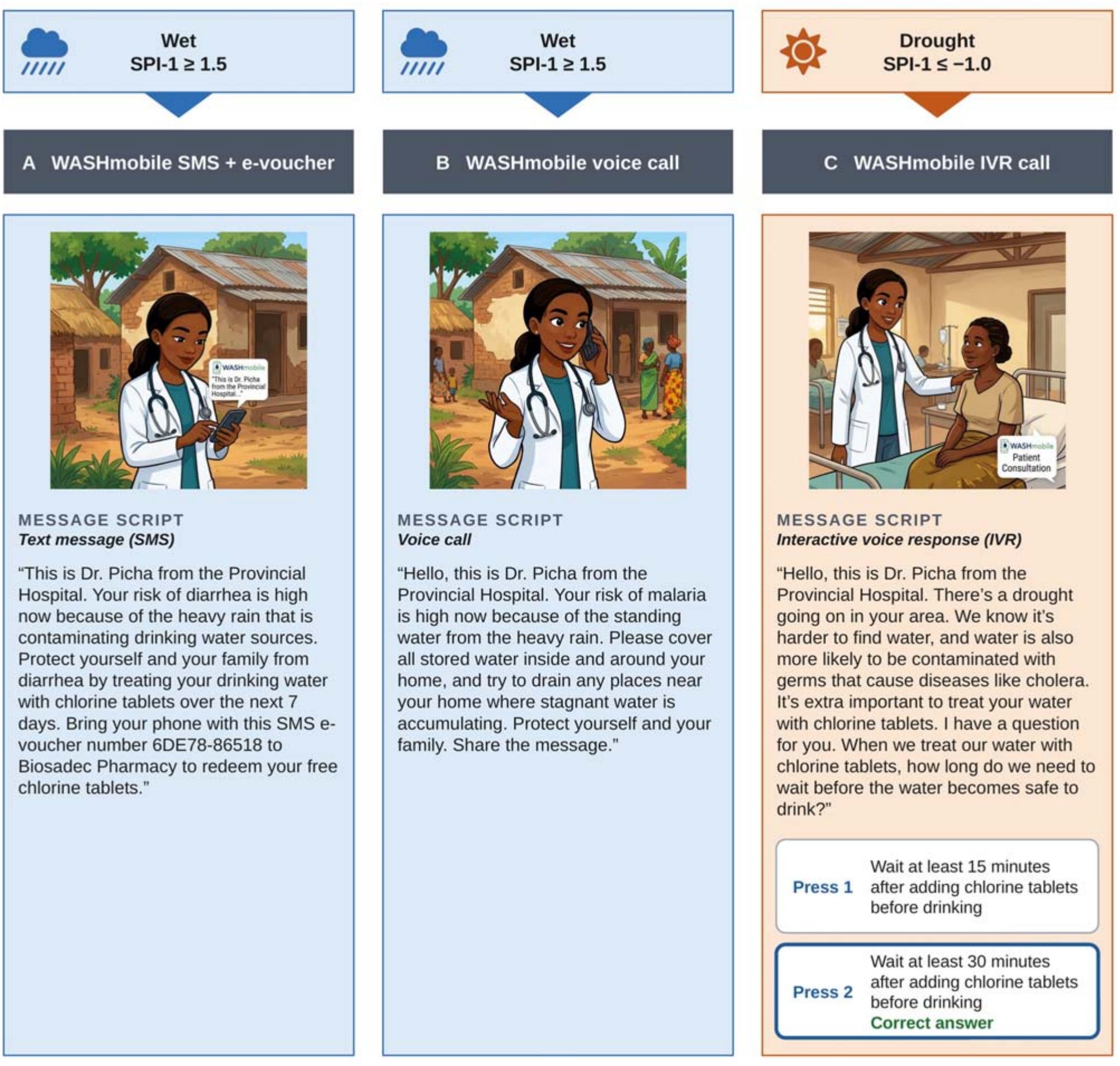
Example mHealth messages to be deployed for early warning and anticipatory action to prevent anemia. When one month of moderate drought, severe wet, or extreme wet conditions is detected, mobile messaging can be deployed through an anticipatory action framework. Example text messages, voice calls, and interactive voice response messages were developed based on expected potential mechanisms for the associations between climate indicators and anemia detected in this study for future WASHmobile deployment.

## DISCUSSION

In this RCT we investigated the association between drought and extreme wet conditions and anemia and assessed the impact of WASHmobile on reducing anemia during these climatic events in a humanitarian context in the DRC. Anemia was significantly associated with moderate drought and extreme and severe wet conditions over the prior month. These associations were consistent across age group and gender with the vast majority of both male and female participants found to have anemia (68%). Furthermore, extreme wet conditions were associated with higher anemia prevalence in the standard arm (78% vs. 62%) but not the WASHmobile arm (64% vs. 69%), which was supported by significant effect modification between extreme wet conditions and WASHmobile delivery on anemia prevalence. Our findings provide evidence to support the potential for WASHmobile, which integrates disease surveillance with targeted WASH, to be implemented as a climate-resilient anticipatory action and early warning approach for anemia prevention during extreme weather.

Several findings from our study suggest that infectious diseases are an important pathway through which extreme weather events influence anemia in our study context. The associations between drought or extreme wet and anemia were observed with one-month SPI but not three-month SPI, which suggests the acute effects of infection. Infection is a major driver of anemia in the DRC, where only 16.5% of anemia has been attributed to iron deficiency.^27^ Diarrhea can cause intestinal inflammation, impaired nutrient absorption, and micronutrient deficiencies,^28^ and malaria infection is commonly linked to anemia.^27^ Extreme wet conditions can contaminate water supplies and increase breeding sites for mosquitoes, while drought can reduce water available for handwashing and push households towards riskier, more contaminated water sources.^5^ Consistent with this, a study nested within the same population observed higher odds of cholera during extreme wet and drought periods (Endres et al. submitted). Food insecurity is also a plausible pathway during severe and extreme wet conditions, as intense rainfall at our study site frequently causes landslides and flooding that destroy crops and disrupt access to markets, and drought can reduce crop productivity. However, these effects would be expected to accumulate over longer periods than the one-month window in which we observed associations.

Our RCT findings suggest that WASHmobile may be effective in reducing anemia during extreme wet conditions. Anemia prevalence was significantly higher during extreme wet conditions compared to near normal conditions in the standard arm but not the WASHmobile arm, with significant effect modification supporting this association. In the parent trial, WASHmobile significantly reduced healthcare facility visits for diarrhea, diarrhea prevalence, cholera serological responses, and stunting for children under 5 years, and led to sustained improvements in handwashing with soap and water treatment throughout the study period.^18^ A plausible pathway is therefore that WASHmobile sustained handwashing with soap and water treatment during extreme wet conditions, preventing enteric infections and thus associated anemia. These findings align with evidence from the WASH Benefits Bangladesh trial, which provided evidence that a WASH intervention contributed to reducing anemia, though the effect of the intervention on anemia was not disaggregated by climate condition.^29,30^ These findings support the potential for WASH programs to be tailored into anticipatory action frameworks to target reductions in anemia during extreme weather.

Our findings led to the development of an anticipatory action framework of evidence-informed climatic triggers to deploy WASHmobile and other WASH interventions to prevent increases in anemia during extreme weather (Figure 4). Because anemia was associated with SPI over the previous month rather than over three months, the window for action is likely short, favoring climatic triggers that can be detected quickly. Additionally, seasonal forecasts can allow time to prepare before the threshold for climatic triggers is met. When an El Niño year is forecasted, for example, anticipatory action such as prepositioning access to chlorine stockpiles could be taken months in advance to reduce the proliferation of enteric and vector-borne diseases. WASHmobile is well suited to this anticipatory action approach because it already activates in response to a case-based trigger indicating the early signs of a diarrhea outbreak,^17^ and mobile message delivery allows extreme weather-specific messages to reach households within hours of a threshold being crossed. The WASHmobile anticipatory approach could also strengthen existing programs, such as those for nutrient and vitamin supplementation, staple food fortification, and education on nutritional diversity. Aligning these activities and WASHmobile delivery with predefined climate triggers could improve effectiveness during drought or extreme wet conditions. Future studies should investigate the implementation of the WASHmobile anticipatory action frame-work in different contexts globally.

This study has several strengths. First, hemoglobin was measured across all ages and sexes, whereas previous studies have focused mostly on women of reproductive age. Second, to our knowledge this is the first study to examine drought and wet conditions in relation to anemia in the DRC, and only the second study of climate conditions and anemia specific to Africa.^10^ Third, the consideration of 1- and 3-month accumulation periods allowed us to consider both acute infection-related and longer-term nutritional mechanisms. Fourth, the RCT component of this study investigated the impact of the WASHmobile program on anemia during extreme weather events through an effect modification analysis. Finally, the findings from this study were used to develop an evidence-informed anticipatory action framework to reduce anemia during extreme weather.

This study has some limitations. First, although the trial was randomized by study arm, the interaction between SPI category and study arm was a post-hoc subgroup analysis that was not prespecified. Therefore, we cannot establish causality about the relationship between WASHmobile delivery and anemia. Second, a point-of-care hemoglobin analyzer was used instead of the gold-standard laboratory-based hemoglobin assessment. Finally, the small geographic study area limits the generalizability of findings to other regions. Future similar studies should be conducted in other contexts globally.

Drought and extreme and severe wet conditions were associated with higher anemia in a humanitarian context in the DRC. Our findings suggest that WASHmobile may present a potential climate-resilient intervention to prevent anemia during periods of extreme wet conditions, likely through reducing infections. These findings highlight the importance of integrating climate-resilient WASH and vector-control interventions into anticipatory action frameworks, with pre-defined climate triggers and early action to prevent anemia before and during extreme weather events. These approaches are critical in humanitarian settings, where anemia prevalence is often high and communities are vulnerable to extreme weather events.

## Data Availability

All data produced in the present study are available upon reasonable request to the authors.

## Contributors

Conceptualization, HG, KE, and CMG; methodology, HG, KE and CMG; software, HG and KE; validation, KE, CMG, JP, LC, AM, AD; formal analysis, HG, KE, CMG, JP; laboratory analysis, AN and WF; investigation, PS, LB, AN, KE, CMG; resources, CMG; data curation, AM, KE; writing—original draft preparation, HG; writing—review and editing, HG, KE, CMG, JP, LC, AD, JB, JCK, JCB, CW, AN, WF, GM, JCC, AM; visualization, HG; supervision, CMG, AM, and KE; project administration, CMG and KE; funding acquisition, CMG. All authors have read and agreed to the published version of the manuscript.

## Institutional Review Board Statement

The study was conducted in accordance with the Declaration of Helsinki, and approved by the Institutional Review Boards of The Catholic University of Bukavu (7107) and the Johns Hopkins Bloomberg School of Public Health Institutional Review Board (9848). The trial is registered at ClinicalTrials.gov (NCT05166850).

## Informed Consent Statement

Informed consent was obtained from all subjects involved in the study.

## Data sharing

De-identified data and the code used for analysis are available from the corresponding author on request.

## Acknowledgments

We thank the participants of this study and all research officers who led the collection of data. We thank all staff of the DRC Ministry of Health in South Kivu, as well as Dr. Placide Welo, Dr. Jean Claude Kulondwa, and Dr. Roger Boketsu.

## AI Use Statement

During the preparation of this work, the authors used Claude for R code development and debugging, Claude for figures, and Gemini for demonstrative images. The authors reviewed and edited the output as needed and take full responsibility for the content of the published article.

## Abbreviations

The following abbreviations are used in this manuscript:

DRC: Democratic Republic of the Congo
CHIRPS: Climate Hazards Group InfraRed Precipitation with Station data
CI: Confidence Interval
GEE: Generalized estimating equation GPS Global Positioning System
IVR: Interactive voice response
RCT: Randomized Controlled Trial
SMS: Short Message Service
WASH: Water, sanitation, and hygiene
SPI: Standardized Precipitation Index
SPI-1: One-month Standardized Precipitation Index
SPI-3: Three-month Standardized Precipitation Index
OR: Odds ratio
WHO: World Health Organization

## Appendix

Appendix Table S1. Prevalence of anemia by age and sex, based on hemoglobin concentrations measured during clinical surveillance in South Kivu, DRC

Appendix Table S2. Association between hemoglobin concentration and climate conditions

Appendix Table S3. Sensitivity analysis: Restriction to standard arm

Appendix Table S4. Associations between climate conditions and anemia status or hemoglobin concentration, stratified by sex

Appendix Table S5. Effect modification by study arm in associations between SPI-1 and anemia.

