## Supplementary File for "Targeted WASHmobile Program for Climate Anticipatory Action to Reduce Anemia during Extreme Weather Events in the Democratic Republic of the Congo"

**Supplementary Table S1:** Prevalence of anemia by age and sex, based on hemoglobin concentrations measured during clinical surveillance in South Kivu, DRC

| **Age group** | Sex | Participants | With anemia | Prevalence |
|---|---|---|---|---|
| <2 years | Male | 80 | 61 | 76.2% |
| | Female | 90 | 70 | 77.8% |
| 2–4 years | Male | 100 | 74 | 74.0% |
| | Female | 102 | 66 | 64.7% |
| 5–11 years | Male | 144 | 95 | 66.0% |
| | Female | 179 | 116 | 64.8% |
| 12–14 years | Male | 30 | 20 | 66.7% |
| | Female | 48 | 34 | 70.8% |
| ≥15 years | Male | 112 | 63 | 56.2% |
| | Female | 323 | 223 | 69.0% |
| **Overall** | **All** | **1208** | **822** | **68.0%** |

Anemia status is presented for 1208 participants, with prevalence reported as the number of participants ever anemic over the study period. Anemia status is characterized according to thresholds set in the 2024 WHO guidelines for determining anemia based on sex and age. Hemoglobin levels are adjusted for altitude of residence using the formula hemoglobin (g/L) − (0.0056384 × elevation + 0.0000003 × elevation$^2$).

**Supplementary Table S2:** Association between hemoglobin concentration and climate conditions

| SPI category | β, g/L | 95% CI | |
|---|---|---|---|
| **SPI-1** | | | |
| Severe/Extreme drought | -0.10 | -4.67 | 4.47 |
| Moderate drought | **-5.69** | **-9.54** | **-1.84** |
| Near normal | | Ref | |
| Moderate wet | -1.46 | -5.01 | 2.08 |
| Severe wet | **-8.87** | **-16.15** | **-1.59** |
| Extreme wet | -1.30 | -7.69 | 5.08 |
| **SPI-3** | | | |
| Severe/Extreme drought | -0.06 | -5.63 | 5.51 |
| Moderate drought | -1.81 | -6.29 | 2.68 |
| Near normal | | Ref | |
| Moderate wet | **3.17** | **0.09** | **6.24** |
| Severe wet | **4.37** | **0.85** | **7.90** |
| Extreme wet | 4.11 | -4.93 | 13.16 |

1520 participant-visits from 1189 participants are included. Participants missing data on any covariates (age, sex, concrete floor, household density, and presence of a household income source) were not included in this model (n=19). Linear models were adjusted for age, sex, study arm, concrete floor, household density, and presence of a household income source, and used generalized estimating equations with clustering by household. Β coefficients for hemoglobin concentration indicate the coefficients for hemoglobin concentration (outcome) on SPI category (predictor), with adjustment for age, sex, study arm, concrete floor, household density, and presence of a household income source. All visits from the standard arm were included, while only the baseline visits of the WASHmobile arm were included in this analysis because the intervention may have altered WASH behaviors associated with infections and, in turn, anemia. Hemoglobin measurements outside the clinically plausible range of 40–200 g/L were removed from analysis as they suggested invalid measurements. CI=confidence interval. Ref=reference category. SPI-1=1-month Standardized Precipitation Index. SPI-3=3-month Standardized Precipitation Index. β=regression coefficient.

Bold = 95% CI excludes the null (β 0).

**Supplementary Table S3:** Sensitivity analysis: Restriction to standard arm

| | Primary analysis sample | | | | | | Standard arm only | | | | | |
|---|---|---|---|---|---|---|---|---|---|---|---|---|
| **SPI category** | n | N | % | OR | 95% CI | | n | N | % | OR | 95% CI | |
| **SPI-1** | | | | | | | | | | | | |
| Severe/Extreme drought | 49 | 73 | 67.1% | 1.23 | 0.73 | 2.05 | 35 | 54 | 64.8% | 1.10 | 0.67 | 1.82 |
| Moderate drought | 113 | 152 | 74.3% | **2.09** | **1.28** | **3.40** | 88 | 116 | 75.9% | **2.32** | **1.31** | **4.10** |
| Near normal | 534 | 916 | 58.3% | | Ref | | 356 | 616 | 57.8% | | Ref | |
| Moderate wet | 184 | 272 | 67.6% | 1.47 | 0.98 | 2.19 | 99 | 137 | 72.3% | 1.78 | 1.00 | 3.16 |
| Severe wet | 29 | 35 | 82.9% | **3.25** | **1.53** | **6.93** | 21 | 27 | 77.8% | **2.43** | **1.24** | **4.79** |
| Extreme wet | 56 | 72 | 77.8% | **2.29** | **1.03** | **5.11** | 56 | 72 | 77.8% | **2.47** | **1.10** | **5.54** |
| **SPI-3** | | | | | | | | | | | | |
| Severe/Extreme drought | 18 | 25 | 72.0% | 1.05 | 0.47 | 2.34 | 11 | 17 | 64.7% | 0.83 | 0.37 | 1.84 |
| Moderate drought | 68 | 94 | 72.3% | 1.27 | 0.80 | 2.01 | 51 | 71 | 71.8% | 1.21 | 0.73 | 2.01 |
| Near normal | 676 | 1051 | 64.3% | | Ref | | 440 | 679 | 64.8% | | Ref | |
| Moderate wet | 78 | 143 | 54.5% | 0.73 | 0.49 | 1.09 | 48 | 87 | 55.2% | 0.70 | 0.43 | 1.14 |
| Severe wet | 108 | 170 | 63.5% | 0.95 | 0.62 | 1.43 | 88 | 131 | 67.2% | 1.09 | 0.68 | 1.74 |
| Extreme wet | 17 | 37 | 45.9% | 0.49 | 0.23 | 1.04 | 17 | 37 | 45.9% | 0.50 | 0.23 | 1.07 |

This sensitivity analysis compares the association between anemia and SPI conditions in the primary analysis sample to a restriction using standard arm visits only. Primary analysis sample: all visits from the standard arm and baseline visits from the WASHmobile arm (1520 participant-visits from 1189 participants). Standard arm only: all standard arm visits (1022 participant-visits from 691 participants). Models for odds ratios used household-clustered logistic GEE with anemia as the outcome and SPI category as the predictor, adjusted for age, sex, concrete floor, household density, and presence of a household income source. Near normal is the reference category for SPI.

CI=confidence interval. GEE=generalized estimating equations. OR=odds ratio. Ref=reference category. SPI-1=1-month Standardized Precipitation Index. SPI-3=3-month Standardized Precipitation Index.

Anemia, n/N (%) and OR (95% CI). Bold = 95% CI excludes the null (OR 1).

**Supplementary Table S4:** Associations between climate conditions and anemia status or hemoglobin concentration, stratified by sex

| | Overall | | | Female | | | Male | | |
|---|---|---|---|---|---|---|---|---|---|
| **Anemia, n/N (%)** | n | N | % | n | N | % | n | N | % |
| **SPI-1** | | | | | | | | | |
| Severe/Extreme drought | 49 | 73 | 67.1% | 25 | 35 | 71.4% | 24 | 38 | 63.2% |
| Moderate drought | 113 | 152 | 74.3% | 69 | 90 | 76.7% | 44 | 62 | 71.0% |
| Near normal | 534 | 916 | 58.3% | 342 | 576 | 59.4% | 192 | 340 | 56.5% |
| Moderate wet | 184 | 272 | 67.6% | 120 | 169 | 71.0% | 64 | 103 | 62.1% |
| Severe wet | 29 | 35 | 82.9% | 17 | 20 | 85.0% | 12 | 15 | 80.0% |
| Extreme wet | 56 | 72 | 77.8% | 29 | 37 | 78.4% | 27 | 35 | 77.1% |
| **SPI-3** | | | | | | | | | |
| Severe/Extreme drought | 18 | 25 | 72.0% | 10 | 12 | 83.3% | 8 | 13 | 61.5% |
| Moderate drought | 68 | 94 | 72.3% | 40 | 53 | 75.5% | 28 | 41 | 68.3% |
| Near normal | 676 | 1051 | 64.3% | 436 | 662 | 65.9% | 240 | 389 | 61.7% |
| Moderate wet | 78 | 143 | 54.5% | 46 | 89 | 51.7% | 32 | 54 | 59.3% |
| Severe wet | 108 | 170 | 63.5% | 62 | 96 | 64.6% | 46 | 74 | 62.2% |
| Extreme wet | 17 | 37 | 45.9% | 8 | 15 | 53.3% | 9 | 22 | 40.9% |

| | Overall | | | Female | | | Male | | |
|---|---|---|---|---|---|---|---|---|---|
| **Odds ratio (95% CI)** | OR | 95% CI | | OR | 95% CI | | OR | 95% CI | |
| **SPI-1** | | | | | | | | | |
| Severe/Extreme drought | 1.23 | 0.73 | 2.05 | 1.30 | 0.57 | 2.98 | 1.19 | 0.67 | 2.10 |
| Moderate drought | **2.09** | **1.28** | **3.40** | **2.13** | **1.25** | **3.62** | **2.25** | **1.09** | **4.63** |
| Near normal | | Ref | | | Ref | | | Ref | |
| Moderate wet | 1.47 | 0.98 | 2.19 | **1.63** | **1.03** | **2.59** | 1.11 | 0.68 | 1.83 |
| Severe wet | **3.25** | **1.53** | **6.93** | 3.09 | 0.92 | 10.38 | **3.53** | **1.17** | **10.68** |
| Extreme wet | **2.29** | **1.03** | **5.11** | 2.28 | 0.79 | 6.64 | **2.72** | **1.06** | **6.95** |
| **SPI-3** | | | | | | | | | |
| Severe/Extreme drought | 1.05 | 0.47 | 2.34 | 1.86 | 0.43 | 8.00 | 0.81 | 0.23 | 2.80 |
| Moderate drought | 1.27 | 0.80 | 2.01 | 1.55 | 0.84 | 2.84 | 1.23 | 0.74 | 2.04 |
| Near normal | | Ref | | | Ref | | | Ref | |
| Moderate wet | 0.73 | 0.49 | 1.09 | **0.62** | **0.39** | **0.97** | 0.87 | 0.45 | 1.69 |
| Severe wet | 0.95 | 0.62 | 1.43 | 0.84 | 0.52 | 1.37 | 1.05 | 0.62 | 1.77 |
| Extreme wet | 0.49 | 0.23 | 1.04 | 0.68 | 0.28 | 1.62 | 0.36 | 0.12 | 1.09 |

| | Overall | | | Female | | | Male | | |
|---|---|---|---|---|---|---|---|---|---|
| **β (95% CI), g/L** | β | 95% CI | | β | 95% CI | | β | 95% CI | |
| **SPI-1** | | | | | | | | | |
| Severe/Extreme drought | -0.10 | -4.67 | 4.47 | -1.24 | -6.28 | 3.79 | 0.88 | -5.01 | 6.78 |
| Moderate drought | **-5.69** | **-9.54** | **-1.84** | **-7.33** | **-11.87** | **-2.80** | **-4.84** | **-9.62** | **-0.05** |
| Near normal | | Ref | | | Ref | | | Ref | |
| Moderate wet | -1.46 | -5.01 | 2.08 | -1.61 | -5.36 | 2.15 | -0.50 | -5.15 | 4.14 |
| Severe wet | **-8.87** | **-16.15** | **-1.59** | -9.05 | -18.77 | 0.66 | **-9.55** | **-16.42** | **-2.68** |
| Extreme wet | -1.30 | -7.69 | 5.08 | -0.31 | -6.03 | 5.40 | -3.63 | -11.75 | 4.48 |
| **SPI-3** | | | | | | | | | |
| Severe/Extreme drought | -0.06 | -5.63 | 5.51 | -1.84 | -6.14 | 2.47 | 2.12 | -5.37 | 9.60 |
| Moderate drought | -1.81 | -6.29 | 2.68 | -4.48 | -9.86 | 0.90 | 0.36 | -5.30 | 6.02 |
| Near normal | | Ref | | | Ref | | | Ref | |
| Moderate wet | **3.17** | **0.09** | **6.24** | **4.79** | **1.34** | **8.24** | 1.08 | -3.51 | 5.68 |
| Severe wet | **4.37** | **0.85** | **7.90** | **5.17** | **1.97** | **8.37** | 4.08 | -0.79 | 8.95 |
| Extreme wet | 4.11 | -4.93 | 13.16 | 5.89 | -0.56 | 12.35 | 3.69 | -8.37 | 15.76 |

1520 participant-visits from 1189 participants are included. Participants missing data on any covariates (age, sex, concrete floor, household density, and presence of a household income source) were not included in this model (n=19). Models were adjusted for age, sex, study arm, concrete floor, household density, and presence of a household income source, and used generalized estimating equations with clustering by household. All visits from the standard arm were included, while only the baseline visits of the WASHmobile arm were included in this analysis because the intervention may have altered WASH behaviors associated with infections and, in turn, anemia. Hemoglobin measurements outside the clinically plausible range of 40–200 g/L were removed from analysis as they suggested invalid measurements.

CI=confidence interval. OR=odds ratio. Ref=reference category. SPI-1=1-month Standardized Precipitation Index. SPI-3=3-month Standardized Precipitation Index. β=regression coefficient.

Bold = 95% CI excludes the null (OR 1; β 0).

**Supplementary Table S5:** Effect modification by study arm in associations between SPI-1 and anemia

| | Anemia, overall | | | Anemia, standard arm | | | Anemia, WASHmobile arm | | | OR (95% CI); p value, SPI-1 category vs near normal, standard arm | | | | OR (95% CI); p value, WASHmobile vs standard arm, near normal | | | | Interaction OR (95% CI); p value, WASHmobile vs standard arm | | | |
|---|---|---|---|---|---|---|---|---|---|---|---|---|---|---|---|---|---|---|---|---|---|
| **SPI-1 category** | n | N | % | n | N | % | n | N | % | OR | 95% CI | | p value | OR | 95% CI | | p value | OR | 95% CI | | p value |
| Severe/Extreme drought | 85 | 112 | 75.9% | 31 | 48 | 64.6% | 54 | 64 | 84.4% | 1.18 | 0.64 | 2.16 | 0.598 | | - | | | 1.64 | 0.50 | 5.32 | 0.414 |
| Moderate drought | 101 | 134 | 75.4% | 57 | 78 | 73.1% | 44 | 56 | 78.6% | **2.12** | **1.04** | **4.32** | 0.038 | | - | | | 0.78 | 0.25 | 2.45 | 0.675 |
| Near normal | 401 | 611 | 65.6% | 192 | 310 | 61.9% | 209 | 301 | 69.4% | | Ref | | | 1.48 | 0.95 | 2.30 | 0.087 | | Ref | | |
| Moderate wet | 79 | 91 | 86.8% | 46 | 53 | 86.8% | 33 | 38 | 86.8% | **3.55** | **1.33** | **9.50** | 0.012 | | - | | | 0.75 | 0.13 | 4.43 | 0.747 |
| Severe wet | 27 | 35 | 77.1% | 12 | 16 | 75.0% | 15 | 19 | 78.9% | 1.92 | 0.69 | 5.35 | 0.213 | | - | | | 0.97 | 0.22 | 4.28 | 0.971 |
| Extreme wet | 119 | 171 | 69.6% | 56 | 72 | 77.8% | 63 | 99 | 63.6% | 3.06 | 1.00 | 9.32 | 0.050 | | - | | | **0.27** | **0.08** | **0.97** | 0.044 |

Hemoglobin levels for all visits after baseline are included, totaling 1154 participant-visits from 945 participants (577 participant-visits from 471 participants in the standard arm, and 577 participant-visits from 474 participants in the WASHmobile arm). Logistic regression for the odds of anemia was performed using generalized estimating equations clustered by household and adjusting for age, sex, and household covariates (presence of a household income source, household density, concrete floor), with a multiplicative interaction term for study arm and SPI category. Study-arm interaction models adjusted for age, sex, people per room, presence of a household income source, and concrete floor. Near normal is the reference category for SPI, and the standard arm is the reference category for study arm. Odds ratios are reported as OR (95% CI).

CI=confidence interval. OR=odds ratio. Ref=reference category. SPI-1=1-month Standardized Precipitation Index.

Bold = $p < 0.05$.
